# USING RE-AIM TO IDENTIFY IMPLEMENTATION DETERMINANTS OF CARDIOVASCULAR RISK STRATIFICATION SCORE FOR PATIENTS WITH PROSTATE CANCER

**DOI:** 10.64898/2025.12.18.25342579

**Authors:** Harikrishnan Hyma Kunhiraman, Aaron J Kruse-Diehr, Phillip J Koo, Pedro Barata, Sagar A Patel, Andrew John Armstrong, Umang Swami, Alicia K. Morgans, Joshi Alumkal, Andrew W. Hahn, Ganesh Palapattu, Zachary Klaassen, Himisha Beltran, Irbaz B. Riaz, Soumyajit Roy, Kelvin Moses, Ashanda R Esdaille, Michael R. Harrison, Neeraj Agarwal, Avirup Guha

**Affiliations:** Division of Cardiology, Department of Medicine, Medical College of Georgia at Augusta University, Augusta, GA 30912; Cardio-Oncology Program, Medical College of Georgia at Augusta University, Augusta, GA 30912; Department of Medicine, Medical College of Georgia at Augusta University, Augusta, GA 30912; Department of Community & Behavioral Health Sciences, School of Public Health, Augusta University, Augusta, GA 30904; Prostate Cancer Foundation, Santa Monica, CA 90401; Clinical Genitourinary Medical Oncology Research Program, UH Seidman Cancer Centre, Cleveland, OH 44106; Department of Radiation Oncology, Emory Winship Cancer Institute Comprehensive Cancer Centre, Emory University, Atlanta, GA 30308; Department of Medicine, Duke University School of Medicine, Durham, NC 27710; Division of Oncology, Department of Internal Medicine, Huntsman Cancer Institute, University of Utah, Salt Lake City, UT, 84132; Department of Medical Oncology, Dana-Farber Cancer Institute, Harvard Medical School, Boston, MA 02215; Genitourinary Medical Oncology Section, Division of Hematology-Oncology, Rogel Cancer Center, University of Michigan Medical School, Ann Arbor, MI 48109; Department of Genitourinary Medical Oncology, Division of Cancer Medicine, The University of Texas MD Anderson Cancer Centre, Houston, TX, 77030; Department of Urology, University of Michigan Medical School, Ann Arbor, MI 48109; Department of Urology, WellStar Medical College of Georgia at Augusta University, Augusta, GA 30912; Department of Medicine, Dana-Farber Cancer Institute, Harvard Medical School, Boston, MA 02215; Department of Hematology and Medical oncology, Mayo Clinic Comprehensive Cancer Center, Phoenix, AZ 85054; Department of Radiation Oncology, RUSH University, Chicago, Illinois 60612; Division of Urologic Oncology, Department of Urology, Vanderbilt University Medical Center, Nashville, TN 37232; Division of Medical Oncology, Duke Cancer Institute, Duke Health, Durham, NC 27710

## Abstract

**Background:** Cardiovascular disease (CVD) remains the leading cause of death among patients with prostate cancer. While the American Heart Association’s PREVENT Score offers a comprehensive lab-based CVD risk prediction model, its integration into oncology workflows is hindered by logistical barriers. To address this gap, the GUHA-STABELLINI Score was developed as a simplified, lab-independent tool tailored for use in specialty care settings.

**Objective:** To evaluate physician preferences, implementation feasibility, and contextual fit of the GUHA-STABELLINI versus PREVENT Score using the RE-AIM (i.e., Reach, Effectiveness, Adoption, Implementation, and Maintenance) model.

**Methods:** A cross-sectional survey was administered to 45 oncology-specialized physicians across academic and community settings. The survey, structured around RE-AIM domains, assessed preferences, implementation perceptions, and perceived effectiveness of each tool. Quantitative responses were analyzed descriptively, and qualitative comments were thematically coded.

**Results:** A significant majority (93%) of respondents preferred the GUHA-STABELLINI Score over PREVENT, citing its ease of use and alignment with clinical workflows. Across RE-AIM domains, GUHA-STABELLINI scored highly in adaptability (71%), cost/resource feasibility (65%), perceived effectiveness (87%), and equity of reach and outcomes (75% and 73%, respectively). Respondents emphasized the tool’s real-time usability, low resource dependency, and ability to facilitate shared decision-making without laboratory input.

**Conclusions:** Despite marginally lower predictive precision, the GUHA-STABELLINI Score demonstrates superior feasibility, reach, and clinical utility within oncology clinics. Findings highlight the importance of implementation-informed design in developing decision support tools. Future research should focus on validating clinical outcomes and expanding use across specialties. The GUHA-STABELLINI Score serves as a model for pragmatic, specialty-integrated preventive care solutions in resource-constrained environments.

**Question:** Among oncology-specialized physicians, does the simplified and laboratory-independent GUHA-STABELLINI cardiovascular risk score offer superior perceived implementability and clinical utility compared with the AHA PREVENT Score for prostate cancer patients on androgen deprivation therapy?

**Findings:** In this cross-sectional survey of 45 physicians, 93% preferred the GUHA-STABELLINI Score over the AHA PREVENT Score. Respondents rated GUHA-STABELLINI as highly adaptable to workflow (71%), cost- and resource-feasible (65%), and likely to be effective (87%) and equitable in reach (75%), despite its slightly lower predictive precision.

**Meaning:** A context-aligned, low-burden cardiovascular risk tool such as GUHA-STABELLINI may achieve greater clinical uptake in oncology settings than more complex laboratory-dependent models, highlighting the central role of implementation-informed design in successful adoption.

## INTRODUCTION

Cardiovascular disease (CVD) remains the leading cause of mortality among men undergoing treatment for prostate cancer.^1,2^ Androgen deprivation therapy (ADT), while effective in reducing cancer progression, significantly increases long-term cardiovascular risk.^3,4^ This risk necessitates structured, evidence-based approaches to stratify patients and guide preventive strategies. In primary care, tools such as the American Heart Association’s PREVENT Score offer robust predictive capabilities. This risk calculator is a novel tool developed to estimate the 10- and 30-year risk of cardiovascular disease (CVD), including heart attack, stroke, and heart failure, in U.S. adults aged 30 to 79. Unlike previous risk assessment models, PREVENT integrates traditional risk factors such as age, sex, blood pressure, and cholesterol with indicators of kidney and metabolic health, including eGFR, HbA1c, and urine albumin-to-creatinine ratio. It can also incorporate a social deprivation index, offering a more holistic and personalized approach to cardiovascular risk prediction. This comprehensive assessment aims to support both clinicians and patients in understanding and managing long-term CVD risk more effectively.^5,6^ However, these tools require lipid profiles (e.g., HDL, LDL) and other laboratory inputs (eGFR), which are often not readily available during urology or oncology visits. This creates a significant barrier to integration into specialty care workflows, where clinicians often have limited time, no access to point-of-care testing, and limited latitude to manage cardiovascular conditions directly.^7^

To address this implementation gap, we developed the GUHA-STABELLINI Score, a streamlined, lab-independent cardiovascular risk stratification tool tailored for use in oncology clinics. ^8^ This tool was designed based on the practical realities faced in specialty settings, prioritizing feasibility, usability, and alignment with non-laboratory data typically collected during a cancer-related consultation. Despite slightly lower predictive accuracy compared to the PREVENT Score, GUHA-STABELLINI’s simplicity may promote greater adoption and broader reach.

Nevertheless, poor fit-to-context (i.e., facilitators and barriers to implementation) can impede the diffusion of interventions and innovations to clinical practice. The RE-AIM (i.e., Reach, Effectiveness, Adoption, Implementation, and Maintenance) model was originally developed to evaluate the public health impact of health promotion interventions^9^ but has since expanded to become one of the most widely used implementation science frameworks.^10^ Although it was originally intended to evaluate implementation outcomes, the RE-AIM framework has also frequently been used in program planning and pre-implementation phases^11–13^ to improve contextual alignment of an intervention, i.e., “beginning with the end in mind.” Using RE-AIM during the pre-implementation phase^14^ of a study can help implementation teams consider various ways in which evidence-based practices (EBPs) can maximize outcomes such as adoption, fidelity to an EBP’s theoretically informed components, and sustainability beyond the active intervention period; it can also be used to adapt EBP components or select implementation strategies hypothesized to improve outcomes that might otherwise be challenging to achieve (e.g., equitable patient reach, consistent provider adoption). (**Figure 1**) Accordingly, we used a RE-AIM-based survey to evaluate the perceived implementability and utility of the PREVENT and GUHA-STABELLINI Scores in preparation for future implementation. In this paper, we describe physicians’ preferences for the GUHA-STABELLINI versus PREVENT scores, outline the demographic and professional composition of respondents, examine the pragmatic challenges and facilitators influencing cardiovascular risk-score adoption in oncology clinics, and demonstrate how RE-AIM implementation outcome domains help elucidate the determinants of successful implementation.

**Figure 1:**
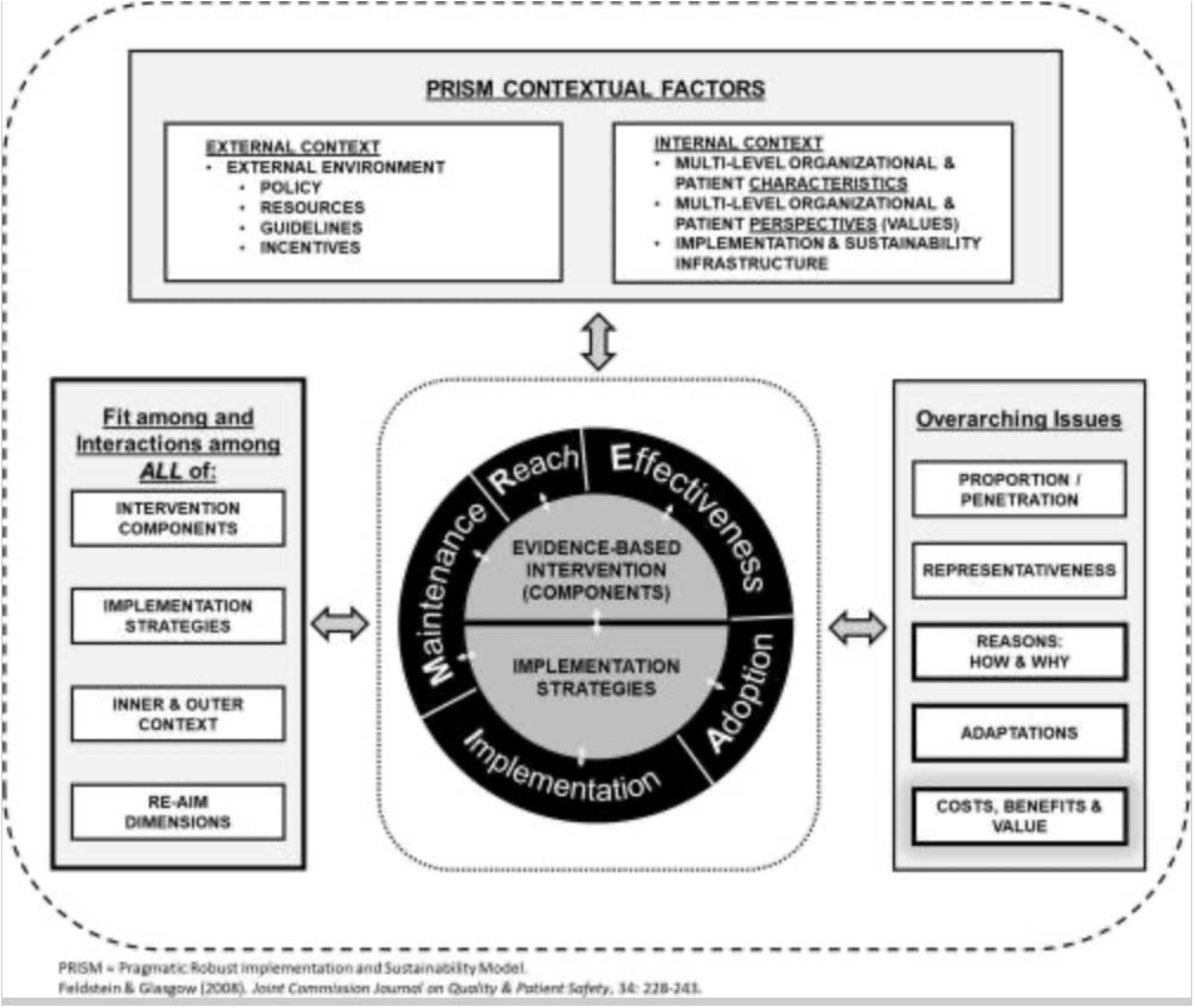
Enhanced RE-AIM Model

- Through this lens, we identify critical themes that can inform future development and scale-up of pragmatic, specialty-specific CVD risk tools and offer evidence for embedding implementation science into the design and evaluation of decision support interventions in oncology.

## METHODS

### Sample and Setting

We developed an educational handout tailored for oncologists, radiation oncologists, and urologists to inform and assess cardiovascular management practices in patients with prostate cancer. Using the ASCO website, we identified and collected contact information for 45 physicians actively involved in prostate cancer care. To minimize potential selection bias, we equitably selected physicians from distinct medical specialties, both academic and community based practice settings, and various geographic regions across the country. We then contacted them via email, sharing the informational pamphlet and requesting their feedback. All 45 physicians consented to participate in the study. After confirming that the content of the pamphlet was well understood, we invited them to complete a brief survey via a Zoom interview. Survey sessions were scheduled individually, and Dr. Guha remained present on each call while participants completed a 12-item multiple-choice questionnaire focused on the implementation of cardiovascular risk calculators in oncology clinics. For added flexibility, an in-person option to complete the survey was also made available during the GU ASCO conference. All survey responses were collected using the Qualtrics platform via a direct survey link.

A total of 45 physicians across oncology-related specialties participated in the cross-sectional survey. The distribution of specialties included medical oncology (48.8%), urology (37.9%), and radiation oncology (13.3%). Most respondents were affiliated with academic institutions (n = 39; 86.7%), while the remainder practiced in private or community-based settings (n = 6; 13.3%). Regarding career stage, 37.8% (n = 17) were late-career physicians with over 20 years of post-residency experience, 35.6% (n = 16) were mid-career with 10–20 years of experience, and 26.7% (n = 12) were early-career with less than 10 years of experience. The sample included 35 male and 10 female physicians. Using the Rural-Urban Commuting Area (RUCA) classification, 95.6% (n = 43) of respondents practiced in urban settings, while 4.4% (n = 2) were in large rural towns. (**Table 1**) No participants were from small rural or isolated areas, underscoring a concentration of oncologic practice in metropolitan and academic environments.

**Table 1:**
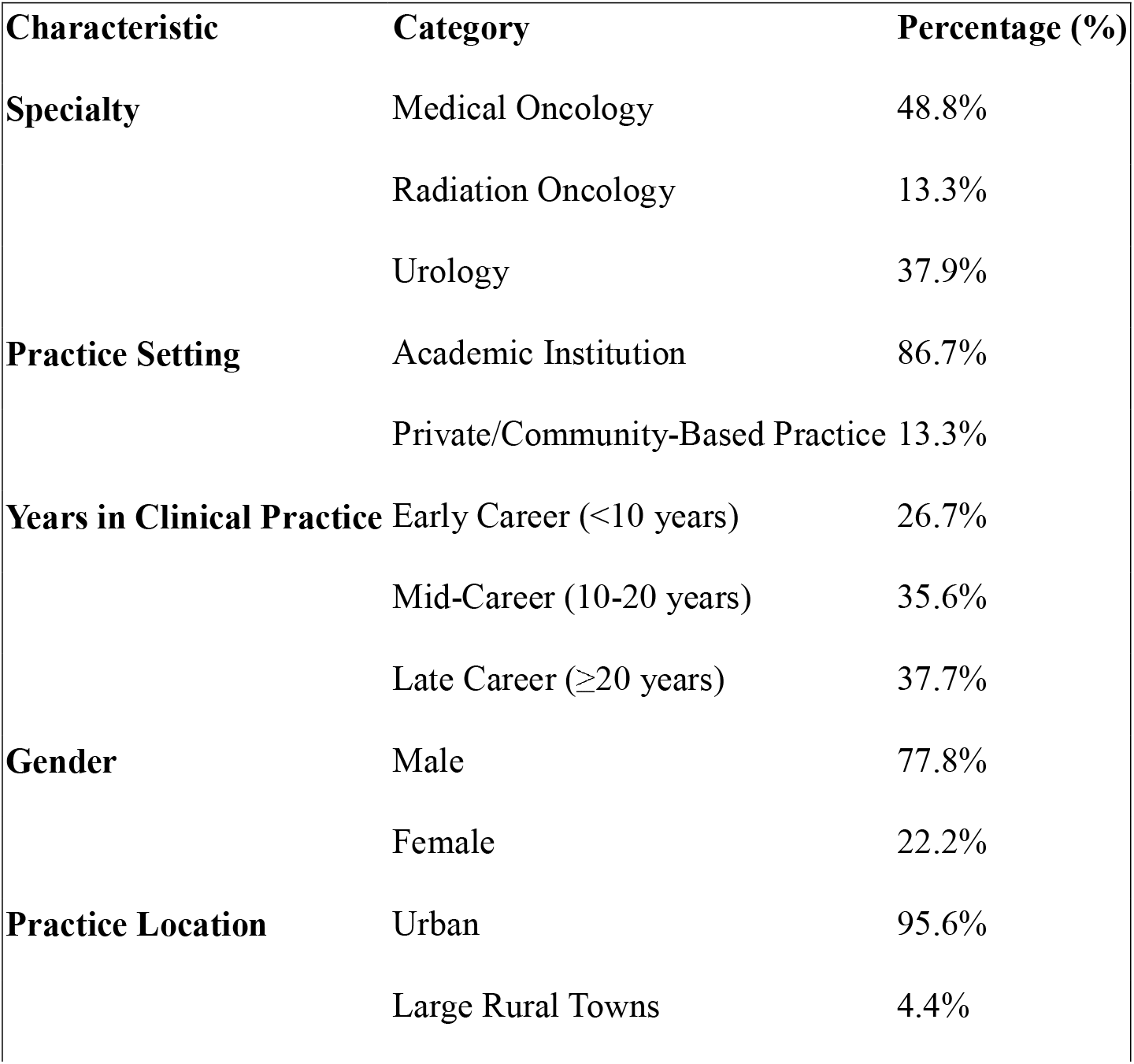
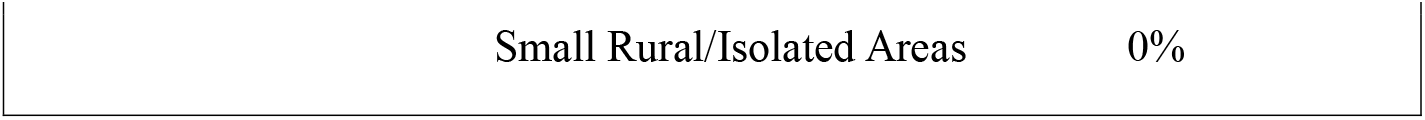
Characteristics of survey takers n=45.

### Survey Instrument

The survey instrument was developed based on constructs from the RE-AIM framework and consisted of three main sections: (1) demographic and professional background questions, (2) a forced-choice item assessing preference between the AHA PREVENT and GUHA-STABELLINI cardiovascular risk calculators, and (3) a series of 11 Likert-scale items designed to evaluate implementation constructs, including likelihood of adoption, perceived effectiveness, equity of outcomes, workflow adaptability, cost feasibility, and long-term sustainability (Appendix B). ^7,11,12^ Finally, participants were invited to write in any comments about the GUHA-STABELLINI cardiovascular risk calculator, risk stratification in general, and foreseeable barriers to implementing the risk calculator; responses to these questions were then mapped to relevant PRISM contextual determinants and RE-AIM outcome domains.

Respondents’ geographic locations were determined via self-reported ZIP codes, which were subsequently analysed for urbanicity using Rural-Urban Commuting Area (RUCA) codes.

### Data Collection and Analysis

This study employed a cross-sectional design using a web-based survey administered through the Qualtrics© platform. The survey was distributed via email to a national sample of physicians practicing in academic and non-academic settings across the United States of America. The recruitment email included a description of the study, an informed consent statement, and a link to the survey. Physicians were eligible to participate if they met the following inclusion criteria: (1) held an MD or DO degree, and (2) were currently practicing in a clinical capacity at the time of survey completion.

## RESULTS

*Preference Analysis:* 42 of the 45 physicians (93%) indicated a preference for the GUHA-STABELLINI Score, with only 3 respondents (7%) preferring the AHA PREVENT Score, further reinforcing the perceived practicality and contextual relevance of the GUHA-STABELLINI tool in specialty settings.

### RE-AIM-Informed Survey Analysis

In addition to overall preferences, 11 targeted survey questions evaluated RE-AIM-aligned implementation outcome domains.^15^ Each used a 5-point Likert scale to quantify feasibility, equity, effectiveness, and sustainability dimensions for the GUHA-STABELLINI score. Regarding adoption of GUHA-STABELLINI, 60% of participants said they were either somewhat or very likely to adopt the risk calculator in routine practice, and 58% responded that clinicians were likely to adopt it equitably across their practices. Importantly, 71% noted that the risk calculator was adaptable to their existing workflows, making it easy for clinicians to implement it with consistency (42% somewhat or very likely). Regarding costs, clinicians responded that GUHA-STABELLINI was somewhat or very likely (65%) to be cost and resource feasible, that it was likely to reach the target population (57%) and achieve equity in its reach (75%). Finally, participants noted that they perceived the risk calculator as likely to be effective (87%), adaptable to their respective practices (69%), and sustainable over time (51%). See **Table 2** for complete response distributions to RE-AIM planning questions. **Error! Reference source not found**.

**Table 2:** RE-AIM-Aligned Survey Questions and Responses.

| Question |  | Physician Responses |  |
| --- | --- | --- | --- |
|  |  | AHA PREVENT CALCULATOR | GUHA-STABELLINI CALCULATOR |
| After reading the Handout, I would prefer the following calculator to be implemented in my clinic |  | 7% | 93% |

| Q/No | Questions | RESPONSES |  |  |  |  |
| --- | --- | --- | --- | --- | --- | --- |
|  |  | Not at all likely | Somewhat unlikely | Neutral | Somewhat likely | Very likely |
| Likelihood of Adoption |  |  |  |  |  |  |
| 1 | How likely is it that your clinic will adopt the Guha-Stabellini Calculator for cardiovascular risk assessment in prostate cancer patients? | 0% | 11% | 29% | 44% | 16% |
| Equity in Adoption |  |  |  |  |  |  |
| 2 | How likely is it that your clinic, regardless of resources or patient demographics, will be able to implement this tool effectively? | 0% | 20% | 22% | 42% | 16% |

| Consistency of Implementation |  |  |  |  |  |  |
| --- | --- | --- | --- | --- | --- | --- |
| 3 | How likely is it that your team will consistently use the calculator with high quality for all relevant patients? | 4% | 13% | 40% | 31% | 11% |
| Adaptability to Workflow |  |  |  |  |  |  |
| 4 | How likely is it that the tool can be adapted to fit the unique workflow of your clinic? | 2% | 7% | 20% | 60% | 11% |
| Cost and Resource Feasibility |  |  |  |  |  |  |
| 5 | How likely is it that the cost and resource requirements for implementing this tool will be feasible for your clinic? | 0% | 16% | 20% | 27% | 38% |
| Reaching in the Intended Patient |  |  |  |  |  |  |
| 6 | How likely is it that the tool will reach the majority of prostate cancer patients in your clinic? | 4% | 13% | 24% | 44% | 13% |
| Equity in patient reach |  |  |  |  |  |  |
| 7 | How likely is it that the tool will equitably benefit all patients, including those from socially and economically disadvantaged backgrounds? | 0% | 7% | 18% | 33% | 42% |
| Perceived Effectiveness |  |  |  |  |  |  |
| 8 | How likely is it that the tool will effectively improve cardiovascular risk stratification in prostate cancer patients? | 0% | 4% | 9% | 38% | 49% |
| Equity in Effectiveness |  |  |  |  |  |  |
| 9 | How likely is it that the tool will be equally effective for patients from diverse socioeconomic and demographic backgrounds? | 0% | 4% | 22% | 33% | 40% |
| Sustainability Over Time |  |  |  |  |  |  |
| 10 | How likely is it that this tool will continue to be used in your clinic over the long term? | 2% | 18% | 29% | 42% | 9% |
| Adaptability Over Time |  |  |  |  |  |  |
| 11 | How likely is it that the tool can be adapted to remain relevant and effective as clinical guidelines and practices evolve? | 2% | 2% | 27% | 47% | 22% |

### Open-Ended Question Themes

Participants overwhelmingly agreed that cardiovascular risk assessment is important for prostate cancer patients on ADT; however, implementation is often fragmented in real-world practice, owing to inconsistency as to who “owns” the responsibility for assessing patient risk (e.g., medical oncology, urology, radiation oncology). As such, clinicians agreed that concrete workflows are necessary to ensure patients are screened consistently. Nevertheless, participants also noted that time pressures make existing workflows challenging to follow consistently, meaning that adoption of the GUHA-STABELLINI score would hinge on delegating workflow tasks, such as utilizing waiting-room time or intake processes and delegating data entry to nursing staff or existing navigation teams. In other words, clinicians are more likely to use the risk calculator if it fits quickly (or invisibly) into their existing workflows. One identified way of achieving this goal is to automate the process in clinic electronic health record (EHR) systems.

Participants provided examples such as automatic data extraction from the EHR and/or prompts that require minimal clicks and do not necessitate manual input of data. Finally, the last major theme noted by clinicians is a need for “next steps”; for example, providers need clear, actionable steps for what to do if the score is high, and they listed examples such as standardized referral recommendations and system-specific pathways (particularly where cardiology access is limited) to ensure their patients receive prompt and responsive care based on their risk stratification.

## DISCUSSION

The findings from this study underscore a critical implementation science insight: clinical effectiveness alone does not guarantee clinical utility. While the PREVENT Score demonstrates superior predictive accuracy due to its integration of lab-based variables like HDL and LDL, its real-world use in oncology and urology clinics is limited by workflow barriers.^7,16–18^ In contrast, the GUHA-STABELLINI Score, though marginally less precise, was favored by the overwhelming majority of physicians due to its pragmatic design.

By applying the RE-AIM framework, we were able to capture multilevel determinants that informed this preference. One of the most salient domains was implementation and sustainability infrastructure.^17^

Oncology clinics, particularly those operating under high patient volume and narrow visit time slots, often lack immediate access to lab values.^16, 19^ The GUHA-STABELLINI Score’s design, requiring no lab work, demonstrated a superior fit within these constraints. This infrastructural compatibility is a key facilitator for sustained adoption.

By looking at the contextual determinant domains of the Practical, Robust Implementation and Sustainability Model (PRISM), a combined determinants-outcomes framework that incorporates RE-AIM domains, we can see how many of the provider responses align with PRISM contextual determinants. For example, participants noted how various *organizational perspectives* might impact implementation: physician engagement with risk stratification tools is contingent on the ability to use these tools during the clinical encounter. The GUHA-STABELLINI Score provided a point-of-care solution that allowed physicians to engage patients in cardiovascular risk discussions without needing to defer or delay until lab results were available.^7,16,18^ Several physicians noted that the tool “enabled shared decision-making in real-time,” which is an important factor in advancing patient-centered care.

Similarly, the PRISM domain of *patient and provider characteristics* also proved revealing. Oncology patients are typically balancing complex therapeutic regimens, emotional distress, and a high volume of information. Risk stratification tools that are cumbersome or require extensive explanation are unlikely to be effective.^7,16,17^ GUHA-STABELLINI was praised for simplifying communication and for aligning with physician judgment, especially among late-career providers who often rely on heuristics and pattern recognition.

Lastly, PRISM’s *external environment* domain can be applied to reveal how systemic challenges like care fragmentation and limited interoperability influence score preference. Physicians in both academic and private settings emphasized that GUHA-STABELLINI’s independence from external data systems, such as centralized lab results or shared EHR platforms, was a major advantage. It minimized delays, avoided unnecessary referrals, and empowered physicians to act within their own clinic’s control.

These findings reflect that the balance between evidence strength and contextual fit is not binary and that it is most always necessary to incorporate relevant implementation strategies into an intervention to support contextual alignment. A slightly less accurate intervention may be more impactful if it overcomes structural and behavioral barriers. As implementation science evolves, more attention must be paid to developing interventions with implementation in mind from the outset, hence the focus of the present pre-implementation study. Our results suggest that pragmatic tools like GUHA-STABELLINI, developed with real-world constraints in mind, can offer significant advantages in clinical adoption. Furthermore, the overwhelmingly positive response across diverse physician groups, including early, mid, and late career, academic vs. private, indicates the generalizability of these insights across settings.

This study also affirms the utility of RE-AIM as a planning framework, in alignment with suggested uses of RE-AIM and PRISM.^20,21^ By “beginning with the ideal outcome in mind” and linking identified barriers with PRISM contextual determinants domains, we are able to consider potential implementation strategies that can address identified barriers and improve fit-to-context in real-world implementation settings. Future studies should consider mixed-method PRISM-informed designs when evaluating implementation readiness of new clinical tools.^14,22^

The expanded data reinforces GUHA-STABELLINI’s strengths in contextual fit. Its successful perception across all RE-AIM dimensions, particularly adaptability (71%), cost feasibility (65%), and perceived effectiveness (87%), strengthens its case as a superior pragmatic tool. Moreover, the data point to GUHA-STABELLINI’s strengths in promoting equity across both adoption and patient outcomes. These insights reflect PRISM’s core values of sustainable, equitable, and patient-centered implementation, especially in resource-variable environments like oncology.

The overwhelmingly high preference rate (93%), coupled with favorable views on implementation feasibility and long-term use, suggest the tool is not only acceptable but potentially transformative in oncology clinics. Respondents emphasized that GUHA-STABELLINI respects workflow constraints and patient realities, key enablers for sustained adoption.

In sum, while the PREVENT Score has stronger empirical foundations, GUHA-STABELLINI offers a better implementation fit for oncology clinics. This highlights the need to balance predictive rigor with contextual feasibility in deploying risk tools across specialty care environments. GUHA-STABELLINI also offers a robust model of how evidence-informed, pragmatically designed tools can successfully integrate into specialty care practices under real-world constraints.

Finally, while this study focused on oncology and urology settings, the principles apply more broadly. Specialty clinics often face similar barriers to integrating general preventive care measures. The GUHA-STABELLINI Score offers a prototype for how simplified, high-utility tools can enable preventive care even in constrained environments.

### Limitations

This study’s modest, predominantly academic sample and its reliance on self-reported perceptions rather than directly observed behaviors may limit generalizability. However, the use of standardized information delivery and balanced representation across oncology specialties enhances the relevance of the findings. Additionally, the presence of the model’s creator during survey administration may have introduced response bias.

### Future Directions

Prospective studies should evaluate patient outcomes from using the GUHA-STABELLINI Score, and comparative-effectiveness research is needed to determine whether its slight reduction in predictive precision is justified by higher adoption rates. Implementation-focused adaptive trials can further refine the tool and identify setting-specific barriers and facilitators.

To address identified barriers, we recommend several implementation strategies to improve workflow integration. EHR integration with automated data pulls, trigger points, and minimal clicks—aligned with ERIC strategies such as *remind clinicians* and *use data-warehouse techniques*—can reduce clinician burden. Delegating data entry and scoring to trained support staff reflects ERIC strategies to *revise professional roles* and *train-the-trainer*. Standardized referral pathways can reduce uncertainty about next steps; ERIC strategies such as *use advisory boards, conduct workshops*, and *stage implementation scale-up* can guide local adaptation and rollout. Provider education, supported by ERIC strategies like *ongoing consultation* and *shadow other experts*, can address knowledge gaps, while patient-facing multilingual materials—aligned with *use mass media* and *provide local technical assistance*—can mitigate language and literacy barriers.

Finally, broader validation across specialties and geographic settings will strengthen external validity and support scale-up. As healthcare systems move toward value-based, preventive care, simple yet sufficiently accurate tools like GUHA-STABELLINI will become increasingly important.

## CONCLUSION

This study highlights how implementation-informed design can drive clinician acceptance of clinical decision support tools, even when those tools offer marginally lower predictive accuracy. While the preference for the GUHA-STABELLINI Score is grounded in its practical advantages, a broader implication emerges: the success of preventive care tools may hinge more on their contextual compatibility than on statistical optimization. Notably, this finding emphasizes the need for clinical innovation to begin with end-user realities, not just epidemiologic data.

Future decision support development should incorporate implementation science frameworks like PRISM from the outset, ensuring tools are not only evidence-based but also feasible, equitable, and sustainable across diverse clinical settings. Additionally, this work signals the potential for simplified tools to serve as bridges between specialty and preventive care, an increasingly important priority in value-based health systems. Widespread uptake of such tools will require continued attention to usability, interoperability, and clinician workflow alignment across healthcare environments.

## Data Availability

All data produced in the present study are available upon reasonable request to the authors

- AG and NA led the development of research questions, study design, and overall scientific direction. Methodology:
- AG, AJKD Designed the analytic strategy, selected the frameworks (e.g., PRISM, RE-AIM, CFIR), and planned statistical/implementation-science methods.
- AG, AJKD, HKHK Assisted in developing analytic pipelines and validated methodological decisions.
- AG Oversaw data procurement, quality checks, harmonization, and cleaning.
- HKHK Verified dataset integrity and resolved discrepancies.
- AG and HKHK performed statistical analyses, quantitative modelling, and interpretation.
- HKHK and AG conducted a literature review, triangulated evidence, and supported data interpretation.
- AG, NA, AJKD provided institutional infrastructure, protected time, equipment, and technical support necessary for the project.
- HKHK and AG wrote the initial manuscript draft, including Introduction, Methods, Results, and Discussion.
- All the other authors reviewed multiple drafts, provided critical intellectual feedback, and approved the final manuscript before submission.

